# Longitudinal Monitoring of Glioblastoma Small Extracellular Vesicle Evolution Using a Nanodiagnostic to Detect Emergence of Glioma Stem Cells Driving Recurrent Disease

**DOI:** 10.1101/2024.09.23.24314250

**Authors:** Zhen Zhang, Richard J Lobb, Paul Tooney, Jing Wang, Rebecca Lane, Quan Zhou, Xueming Niu, Sam Faulkner, Bryan W Day, Simon Puttick, Stephen Rose, Mike Fay, Matt Trau

**Author notes:** Corresponding authors: Richard Lobb; Matt Trau. These authors contributed equally to this work (Zhen Zhang, Richard J Lobb).

## Abstract

Assessing therapeutic response in glioblastoma (GBM) is a major factor limiting the clinical development of new and effective therapies. The intracranial location limits serial biopsies, and only provides an intermittent view of the tumor molecular profile from the initial resection. Liquid biopsy techniques, specifically small extracellular vesicle (sEV) analysis, have the potential to overcome these limitations by providing a window into the brain using peripheral blood. To address the need for monitoring tumor evolution and therapeutic resistance, we developed a GBM biomarker panel (ATP1B2, EAAT2, CD24, CD44, CD133 and EGFR) for multiplexed profiling of sEVs using an advanced GBM Extracellular vesicle Monitoring Phenotypic Analyzer Chip (GEMPAC). We successfully tracked patient response to treatment by monitoring changes in glioma stem cell markers on circulating sEVs. We propose that these results provide a strong rationale for using GBM sEVs as a serial monitoring tool in the future clinical management of GBM patients.

## INTRODUCTION

Glioblastoma (GBM) is an aggressive and invasive primary brain tumor originating in the central nervous system (CNS).[1–4] The prognosis of GBM is very poor, with a median survival time of approximately 14 months.[5] The rapid development of treatment resistance and frequent local recurrence are key features contributing to poor patient outcomes.[1, 2, 4] Accurate monitoring of GBM tumor progression is difficult and expensive; depending on imaging modalities, such as magnetic resonance imaging (MRI).[6] The standard diagnostic methods in GBM management (i.e. histopathology of the resection specimens) provide a limited temporal molecular profile of the tumor and are unable to monitor tumor progression dynamically.[7] In addition, detecting early signs of tumor progression is challenging when using MRI, as the interpretation of diagnostic results may be complicated by pseudo-progression after diagnosis (in the first three months after radiotherapy), and by radiation necrosis (3-12 months after radiotherapy).[8, 9] Therefore, developing non-invasive methods for monitoring of tumor evolution during therapy is essential to navigating treatment management in this aggressive disease.

In recent years, liquid biopsy has emerged as a diagnostic concept that characterizes and monitors tumor signatures in patient blood for tumor subtyping, prognosis monitoring, and assessing treatment response.[10, 11] These non-invasive approaches can provide a view of the molecular landscape of tumors from blood samples.[12] As a liquid biopsy marker, exosomes or small extracellular vesicles (sEVs) are nano-sized vesicles (50-150 nm) released from cells.[7, 13, 14] These vesicles are critical mediators of cell-cell communication and carry molecular cargo, including proteins, nucleic acids, metabolites, and lipids.[7, 13] Most critically, several studies have indicated that sEVs released by GBM tumors can cross the intact blood-brain barrier (BBB) and gain access to the peripheral blood.[15–18] Several studies have reported an overall increase in bulk sEV concentration in GBM patients relative to healthy controls, with sEV load apparently decreasing following successful treatment of the primary tumor.[7, 15, 19, 20] Other studies, however, have suggested that systemic increases in bulk sEV concentration are not a GBM-specific phenomenon, meaning sEV concentration alone is unlikely to be a useful metric.[21, 22] Further work has demonstrated that sEVs may reflect the molecular profile of the GBM primary tumor and provide clinical information to direct treatment in a timely manner[23], thereby providing an opportunity to utilize GBM sEVs as a tool to inform clinical management decisions.

Progress in utilizing GBM sEVs has been limited by the challenge of specifically isolating and characterizing GBM sEVs from non-target bulk sEVs in blood. Previous work has attempted to identify specific protein cargo associated with circulating GBM sEVs, including transmembrane L1 cell adhesion molecule,[24] epidermal growth factor receptor variant III (EGFRVIII) [23], von Willebrand factor [20] and syndecan-1.[25] However, these biomarkers are also expressed in a variety of normal tissues and are unable to specifically isolate GBM sEVs.[24, 26, 27] To address the lack of specificity for interrogating sEVs derived from GBM tissue, we identified a unique GBM signature on sEVs in blood. By integrating a GBM sEV signature with a nanoshearing multiplex surface-enhanced Raman spectroscopy (SERS) platform, termed GBM EV Monitoring Phenotypic Analyzer Chip (GEMPAC), we have developed the capability to sensitively profiling the surface protein composition of sEVs in GBM patients with high precision.

In the study, the GEMPAC assay precisely captures GBM sEVs by targeting our unique CNS signature composed of transmembrane proteins sodium/potassium-transporting ATPase subunit beta-2 (ATP1B2) and excitatory amino acid transporter 2 (EAAT2), both of which are highly expressed in both normal CNS and GBM tissue.[28–30] The malignant phenotype of captured CNS sEVs were profiled by measuring glioma stem cell (GSC) markers to detect the emergence of GSC subpopulations that could potentially drive recurrent disease and therapy resistance.[31] Previous single-cell RNA sequencing data has revealed certain surface markers that characterize cell-likes states of GSCs. In particular, CD24 expression is enriched in neural progenitor cell-like (NPC-like) cells, CD44 in mesenchymal cell-like (MES-like) cells, CD133 in oligodendrocyte-progenitor cell-like (OPC-like) cells, and EGFR in astrocyte-like (AC-like) cells.[32, 33] Using our GEMPAC platform we captured GBM sEVs and monitored GSC subpopulation evolution in patients before and during therapy. Encouragingly, by analyzing GSC subpopulation evolution during therapy, we detected treatment response and tumor progression, thereby opening a new window to facilitate clinical management of GBM. We believe that our diagnostic platform has the potential to facilitate future therapies through monitoring circulating GBM GSC-related sEVs to improve clinical decision-making and therefore patient survival.

## RESULTS

### GEMPAC chip dynamically monitors GBM sEV profiles

To accurately monitor GBM progression, we hypothesized that circulating sEVs derived from the blood of GBM patients carry specific protein profiles that indicate the degree of tumor burden/or progression in the CNS (Figure 1A). Specifically, we designed a panel of six surface proteins capable of capturing CNS-derived sEVs and monitoring GBM by detecting GSC biomarker expression (Figure 1B). We synthesized unique SERS nanotags conjugated with antibodies targeting GSC surface proteins and dedicated Raman molecule reporters on gold nanoparticles bearing distinct SERS signatures indicating GSC protein expression (Supplementary, Figure S1). The sum of GSC protein expression was defined as the GEMPAC score and used to evaluate disease progression (Figure 1C). Using this multiplexed approach, we combined these biomarkers and investigated their capability as a dynamic monitoring tool of GBM in the blood, including monitoring therapy response and detecting tumor progression for improved clinical management (Figure 1C).

**Fig. 1.**
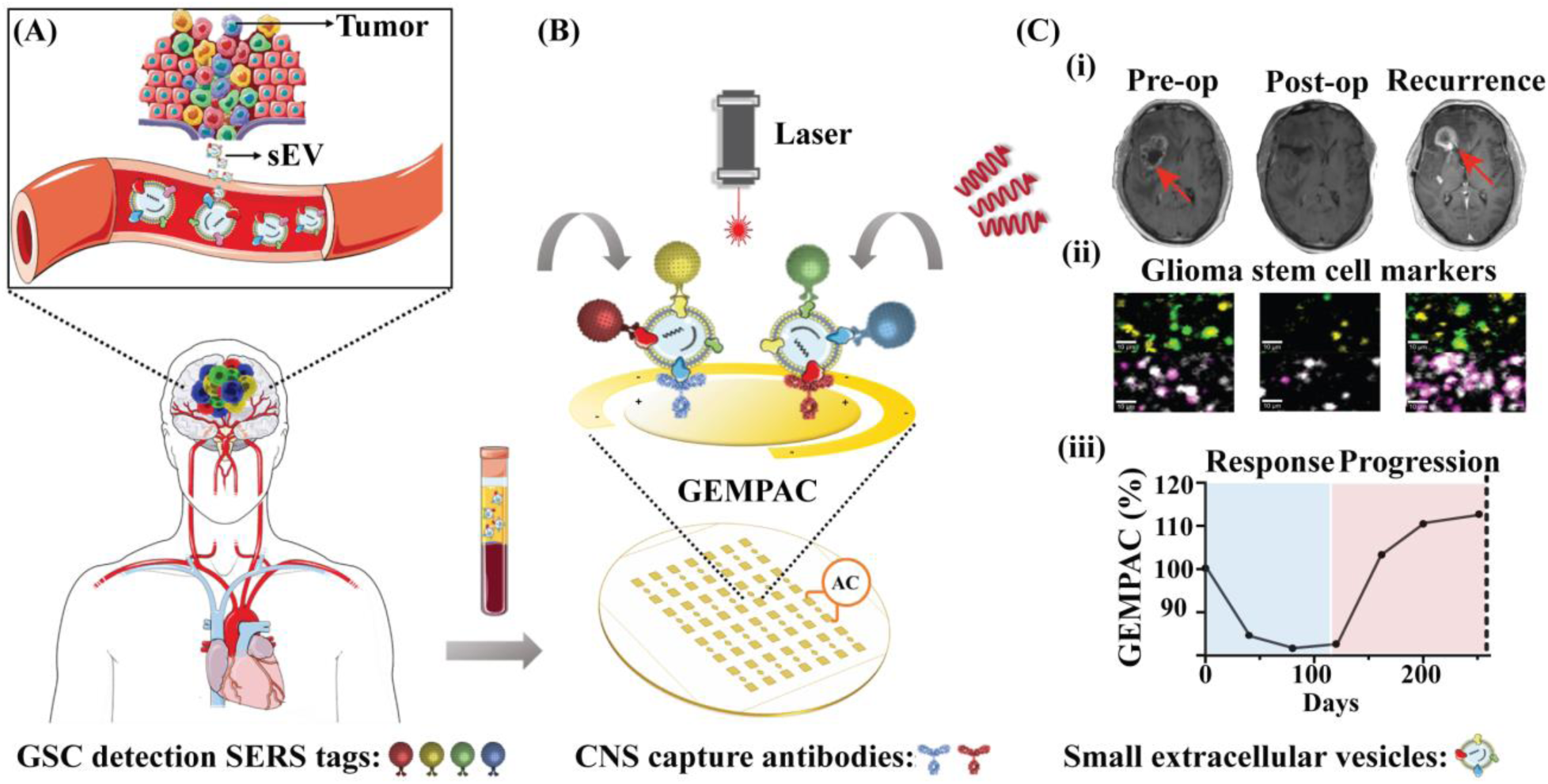
Multiplexed detection of GBM sEVs by the GEMPAC. (A) GBM tumor cells release sEVs into the bloodstream. GBM sEVs cross the BBB into the peripheral circulation. (B) GBM sEVs are purified from blood and enriched using GEMPAC functionalized with CNS capture antibodies for selective capture of CNS sEVs. The GEMPAC platform utilizes an alternating current electro-hydrodynamically (AC-EHD)-induced nanoshearing strategy to minimize nonspecific interactions[34] and improve specific capture of CNS sEVs. After that, SERS nanotags that are conjugated with Raman reporters and detection antibodies are applied to the system for multiplex readout. The SERS nanotags with special Raman molecules include, 5,5’-dithiobis (2-nitrobenzoic) (DTNB) (red), 4-Mercaptobenzoic acid (MBA) (blue), 4-mercaptopyridine (MPY) (yellow) and 2,3,5,6-tetrafluoro-MBA (TFMBA) (green). (C) (i) Representative MRI images for tumor scan tumor at pre-operation (pre-op), post-operation (post-op), and recurrence. (ii) Representative pseudo-colored SERS images indicate the presence of each GSC marker in sEVs. (iii) The changes of GSC score on GBM sEVs in response to the treatment and disease progression. The figure was partly generated using Servier Medical Art.

### Biomarker selection and validation

To capture GBM sEVs in peripheral blood, targeting CNS-related cell surface markers in sEVs is an essential component of this study. CNS markers were selected through bioinformatic analysis of the Genotype-Tissue expression (GTEx) and The Cancer Genome Atlas (TCGA) databases, with cross-reference to all annotated membrane proteins in the UniProt database. Utilizing this approach, we developed a candidate list of membrane protein-coding genes (Figure 2A; Supplementary, Figure S2 – complete heatmap) of which we selected ATP1B2, the β2 subunit of Na (+)/K (+) – ATPase expressed on glial cells[29], and solute carrier family 1 member 2 (SLC1A2), the astrocyte glutamate transporter EAAT2 for further analysis. [35, 36] These markers were selected based on the suitability of appropriate antibodies targeting extracellular membrane domains. Importantly, the RNA expression levels of ATP1B2 and EAAT2 are highly expressed in brain tissue as well as low grade glioma (LGG) and high-grade GBM, compared to other normal tissues and cancers (Figure 2B; Supplementary, Figure S3). Given the hypothesis that sEVs represent their cell-of-origin[23, 37], this differential expression underpins our approach to capture limited GBM sEVs circulating in the blood.

**Fig. 2.**
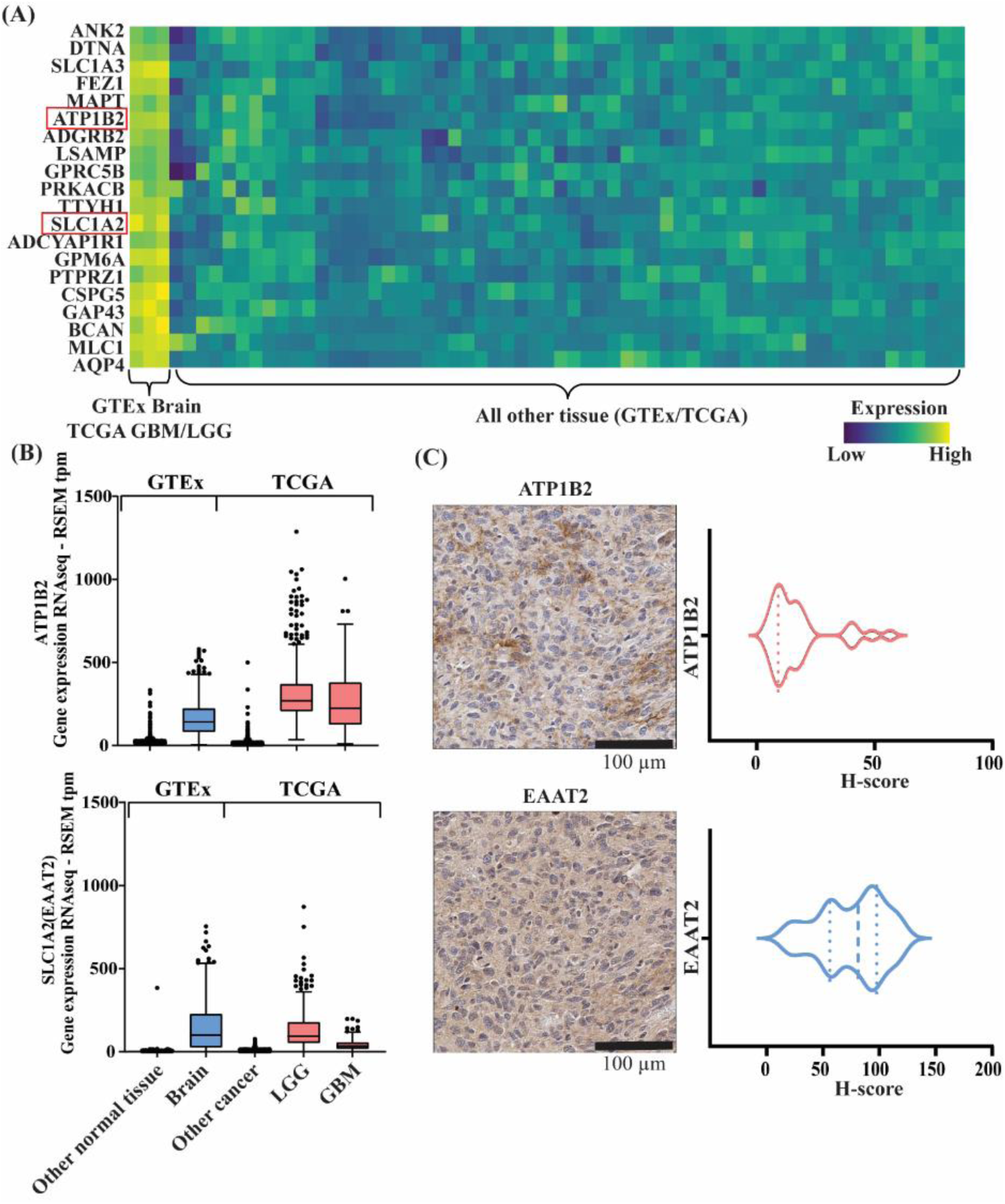
CNS marker selection and validation. (A) Heatmap of top 20 CNS markers’ RNA expression in TCGA and GTEx RNA-seq databases. Low expression is indicated in blue and high expression is indicated in yellow. (B) Tukey’s boxplot indicating *ATP1B2* and *SLC1A2* (EAAT2) genes are highly expressed in normal brain and brain tumor tissues (GBM and low-grade gliomas, LGG) in TCGA and GTEx RNA-seq databases. (C) Primary GBM tumors (n = 31) were labeled for ATP1B2 and EAAT2. Violin plots show the H-score labeling intensity across the cohort. Scale bar = 100 µm.

We then clinically validated our bioinformatic selection of ATP1B2, EAAT2 through IHC of primary GBM tissues. For ATP1B2, low-intensity labeling reflected in low H-scores (mean 17.58 +/- 13.42; range 3.96 - 56.53) was detected across the cohort of tumors (Figure 2C). ATP1B2 labeling was observed diffusely spread throughout the dense neuropil, which normally contains the nerve fibers and synapses and glial processes (Figure 2C). The labeling of ATP1B2 on control tissues was also analyzed, in which strong ATP1B2 labeling of the cell membrane and cytoplasm of photoreceptor cells; cells of the inner nuclear layer of the human retina; and positive labeling of neuropil in human cortex (Supplementary, Figure S4A & B). Moderate EAAT2 labeling was detected in tumor tissue for all cases of GBM (mean 75.89 +/- 28.88; range 18.13 – 119.69; Figure 2C). EAAT2 labeling was detected diffusely throughout the neuropil across the tumor sections with occasional intense labeling of cell membranes and processes (Figure 2C). In the control tissues, no EAAT2 expression were present in colonic crypts but in the human brain cortex strong EAAT2 labeling was observed in astrocytes (Supplementary, Figure S4C & D).

After validating the selection of CNS markers, CD24, CD44, CD133 and EGFR were selected to monitor different cell-like states of GSCs.[31] Specifically, scRNA-seq data has revealed that CD24 is enriched in NPC-like GSCs, CD44 in MES-like GSCs, CD133 in OPC-like GSCs, and EGFR is enriched in AC-like GSCs.[32, 38, 39] These surface GSC markers correlate with an aggressive phenotype, poor survival, and associated with the rapid development of treatment resistance.[31, 40–48] Transcriptomic expression based on the TCGA dataset confirmed that all these markers are expressed in GBM (Supplementary, Figure S5). We then validated the expression of CD44, CD133 and EGFR in GBM tumor tissues through IHC labeling (Supplementary, Figure S4 & S6). EGFR (mean 173.01 ± 54.25; range 2.65 – 259.59) and CD44 (mean 130.58 ± 43.84; range 57.66 – 205.24) were highly expressed across all the tumor tissues with intense labeling of tumor cell membranes. Minimal labeling was detected in most GBM tumor sections for CD133 (mean 0.26 ± 0.80; range 0 – 4.20), with occasional positive labeling detected sparsely throughout the tumor (Supplementary, Figure S6).[45] IHC labelling for CD24 was not completed across the cohort of tumor tissues in this study, as the labelling of the cells and tissue structures within the control tissues with several antibodies was not consistent with that reported in the Human Protein Atlas. However, we chose to continue with CD24 in this study due to the protein expression of GBM tissue in the Human Protein Atlas. Given the expression of these biomarkers in GBM tissues based on TCGA dataset and IHC results, measuring the expression changes of CD24, CD44, CD133 and EGFR on CNS derived sEVs from the same GBM patients may help predict and track disease progression and treatment response.

### Single nanoparticle analysis and GEMPAC marker validation

We then sought to validate our hypothesis that ATP1B2 and EAAT2 are specifically enriched on the surface of GBM sEVs. To establish an *in vitro* model for GBM sEV analysis, the expression of ATP1B2 and EAAT2 in three unique patient-derived GBM cell lines (BAH1, WK1, and FPW1; representing classical or mesenchymal GBM subtypes[49]) were validated using flow cytometry. In support of our bioinformatic selection of CNS candidates and IHC results, ATP1B2 and EAAT2 were highly expressed on the cell surface of three patient-derived GBM cell lines (Supplementary, Figure S3C & D), and therefore as a result have the potential to be present on the surface of GBM sEVs.

Next, we collected sEVs from our GBM patient-derived cell line panels and purified them through size exclusion chromatography (SEC).[50] Purified sEVs were characterized by nanoparticle tracking analysis (NTA), showing a characteristic modal size of 90-100 nm (Supplementary, Figure S7A). We then used transmission electron microscopy (TEM) to visualize the morphology of purified sEVs (Supplementary, Figure S7B). Following this, the surface expression of the canonical sEV tetraspanin CD9 and CD63 were measured by nanoflow cytometry (nanoFCM) (Supplementary, Figure S7C), and western blot demonstrated the presence of internal HSP70 and absence of the endoplasmic reticulum–associated molecular chaperone, calnexin (Supplementary, Figure S7D).

We next evaluated if the expression of ATP1B2 and EAAT2 was specifically enriched on GBM sEVs. As we hypothesized, cell-of-origin-specific enrichment of ATP1B2 and EAAT2 were detected on GBM sEVs (14.22% and 32.37% respectively), while MDA-MB-231-derived breast cancer sEVs (-ve) contained negligible levels of both CNS markers (Figure 3A; Supplementary, Figure S8). Next, we evaluated the expression of the GSC markers CD24, CD44, CD133, and EGFR on the surface of GBM sEVs. Using single nanoparticle analysis by nanoFCM, we found high levels of CD24 (19.1%) and EGFR (23.6%), moderate levels of CD44 (8.17%) and low levels of CD133 (4.06%) (Figure 3B; Supplementary, Figure S8).

**Fig. 3.**
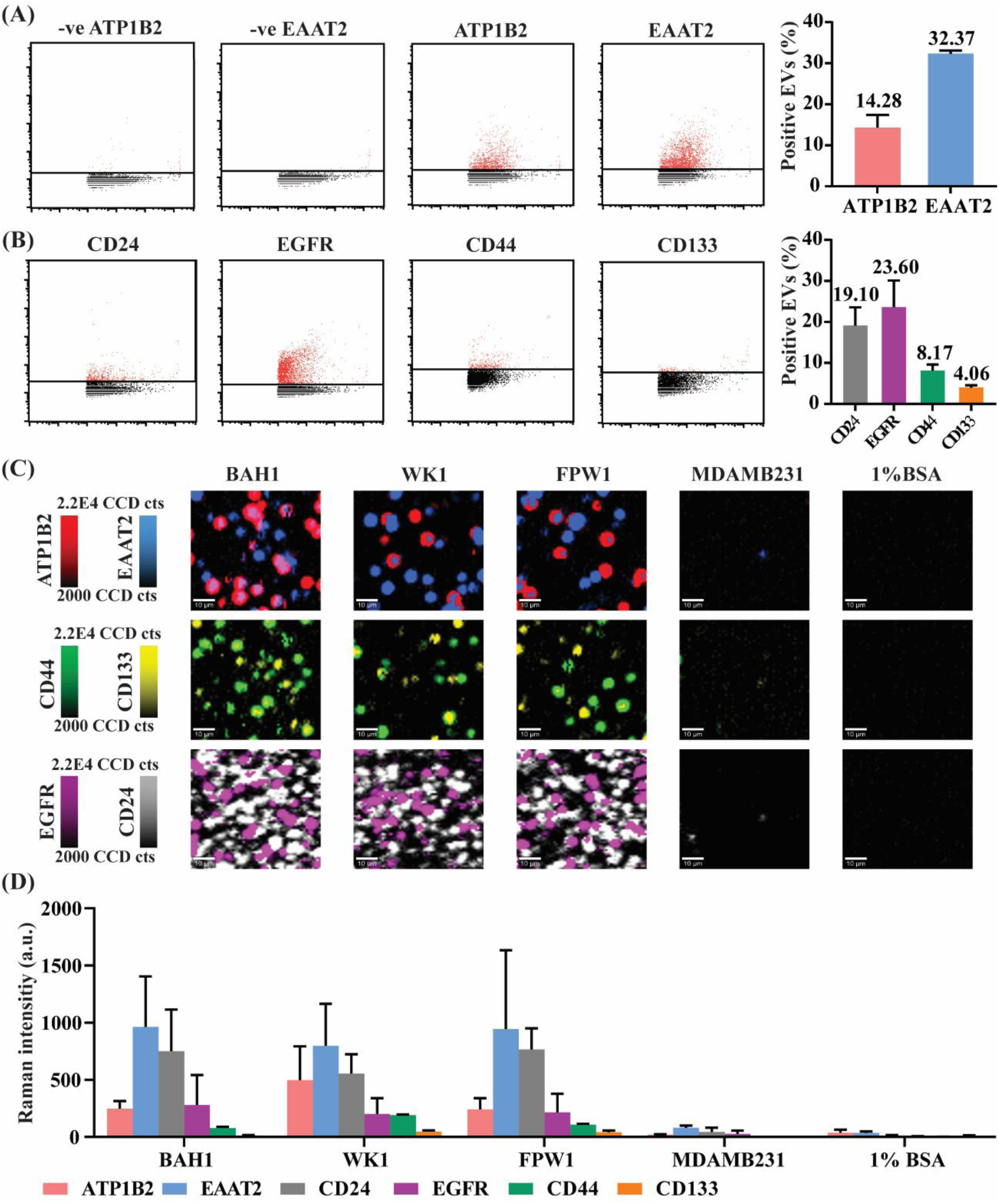
Expression of selected markers on GBM sEVs by single nanoparticle analysis and GEMPAC. (A) Single nanoparticle analysis by nanoFCM shows the expression of CNS markers (ATP1B2 and EAAT2) in breast cancer cell line MDAMB231 (-ve) and WK1 sEVs. Average histogram measurements are the average expression of 3 GBM cell line sEVs (WK1, BAH1, FPW1). (B) Single nanoparticle analysis by nanoFCM shows the expression of GSC markers (CD24, EGFR, CD44, and CD133) as representative nanoFCM images and average histogram measurements in three GBM cell lines. (C) Representative Raman images for all of marker’s detection in GBM cell lines using GEMPAC. Scale bar = 10 µm. (D) Raman intensity measurements of sEVs from each GBM cell line with different SERS nanotags; DTNB-ATP1B2 (red), MBA-EAAT2 (blue), TFMBA-CD44 (green), MPY-CD133 (yellow), TFMBA-EGFR (purple) and MPY-CD24 (grey). MDA-MB-231 breast cancer cells and EV-free medium were used as negative controls to show biomarker specificity. Data are represented as mean ± standard error.

After validating that we have identified a unique CNS signature for the analysis of GBM sEVs, we sought to develop an approach that meets the requirements for sensitive and specific detection of GBM sEVs that is clinically translatable (GEMPAC). We carried out a specificity assay to examine the functionality and specificity of the GEMPAC platform. Each electrode of the GEMPAC was functionalized with dual capture antibodies, anti-ATP1B2 and anti-EAAT2, for capturing GBM sEVs. A purified total of 5 × 10^8^ sEVs particles were then applied to individual electrodes and subsequent AC-EHD nanomixing facilitated specific binding of GBM sEVs while simultaneously reducing non-specific interactions.[34] Following this, unique SERS nanotags were applied to each electrode to detect targeted markers on captured sEVs simultaneously. Pseudo-colored SERS mapping images for GBM sEVs are shown in Figure 3C. High expression of both ATP1B2 and EAAT2 was observed for the GBM cell line-derived sEVs (BAH1, WK1, and FPW1), with minimal signals detected for non-GBM sEVs (MDA-MB-231 breast cancer) and an sEV-free control (Figure 3D). Expression of the putative prognostic GSC markers CD24, CD44, CD133 and EGFR were also detectable in the GBM sEVs and absent from other samples due to the specific capture of GBM sEVs (Figure 3D). As a result, combining these markers could distinguish GBM sEVs from other malignancies.

### Analysis of GBM patient sEVs sampled before and after surgical resection by GEMPAC

Given the specificity of the GEMPAC assay for analyzing GBM sEVs, we next sought to investigate the composition of GBM sEVs in the blood of GBM patients. Pre- and post-op blood plasma were sourced from GBM patients. sEVs were purified from plasma using SEC and analyzed with TEM and nanoFCM to confirm size distribution and concentration of particles (Figure 4A & B). There were no significant differences in sEV size and concentration at the two time points (p > 0.05) (Figure 4B), indicating that these specific sEV properties provide little prognostic or diagnostic information overall. Therefore, our approach of profiling the phenotype of GBM sEVs using GEMPAC may offer more insights for diagnostic and prognostic applications in GBM patients.

**Fig. 4.**
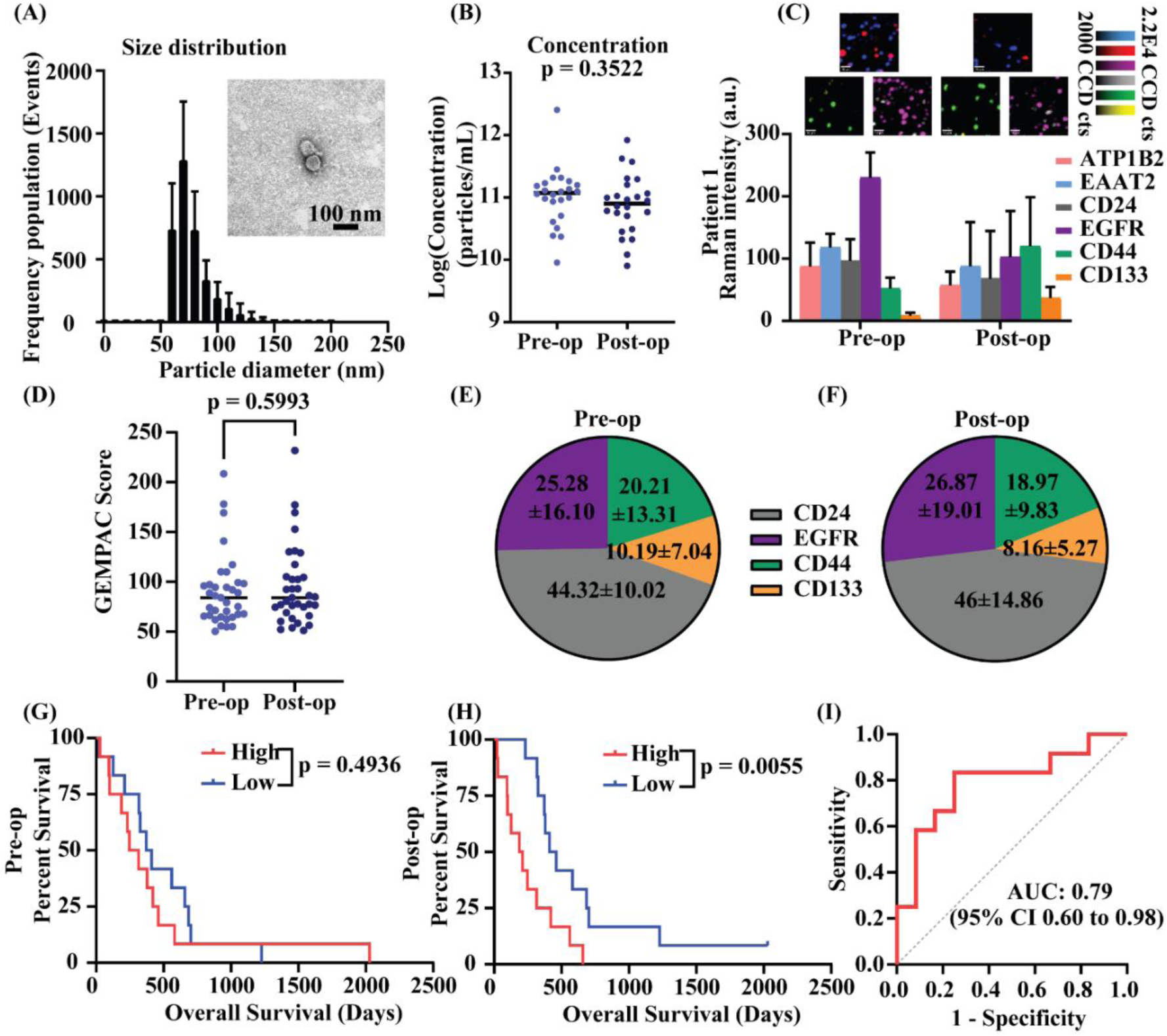
Analysis of sEVs in plasma before and after surgical resection in GBM patients. (A) GBM patient samples (n = 36) sEV size distribution. (B) GBM sEV concentration in pre-op and post-op. (C) Raman intensity measurements of each marker with SERS images in pre-op and post-op in the representative GBM patient, DTNB-ATP1B2 (red), MBA-EAAT2 (blue), TFMBA-CD44 (green), MPY-CD133 (yellow), TFMBA-EGFR (purple), and MPY-CD24 (grey). Data are represented as mean ± standard error (n = 4). (D) All the GBM stem cell markers were normalized by CNS markers and sum as GEMPAC score. Comparison of GEMPAC score between pre-op and post-op. The proportion of each normalized GSC marker in (E) pre-op and (F) post-op. Kaplan-Meier plots of overall survival by GEMPAC score. Survival outcome differences in pre-op (G) and post-op (H). (I) ROC curve analysis for GEMPAC score between short and long overall survival groups. Next, we investigated if the GEMPAC score was able to predict overall survival in a cohort of 24 patients who received only standard care temozolomide (TMZ) and radiotherapy (RT). Patients were categorized into either high (> median GEMPAC score) or low (< median GEMPAC score) groups. Before surgical resection there was no correlation in overall survival (p = 0.4936) (Figure 4G), however, after surgical resection, patients with a high GEMPAC score had significantly shorter survival compared to patients with a low GEMPAC score (p = 0.0055) (Figure 4H). To investigate the sensitivity and specificity of the GEMPAC score for predicting overall survival, a 1-year overall survival was assessed by receiver operating characteristic (ROC) curves. In this context, the area under the curve (AUC) for predicting overall survival after surgical resection (post-op) was 0.79 (95% CI 0.6-0.98) (Figure 4I). These results indicate the GEMPAC score from circulating GBM sEVs could predict patients’ survival after surgery, likely reflecting the remaining tumor burden after surgical resection which is a predictor of survival after resection.[51]

To address this, we successfully profiled GBM sEVs with our GEMPAC platform by quantifying the composition of ATP1B2, EAAT2, CD24 CD44, CD133, and EGFR (Representative patient, Figure 4C). To understand changes in circulating GBM sEVs, each GSC marker was normalized by the CNS markers (ATP1B2 and EAAT2) and combined into the GEMPAC score to provide insights into the tumor burden of GBM patients. A total of 36 patients were profiled to investigate if the GEMPAC score changed after surgical resection. There was no significant change in the GEMPAC score before and after the surgery (p > 0.05) (Figure 4D), and the phenotypic composition of GSC markers on circulating GBM sEVs remained the same (Figure 4E & F).

### Longitudinally monitoring radiological recurrence and sEV evolution by GEMPAC

Our understanding of glioblastoma heterogeneity is limited in terms of the cellular landscape at various stages of disease evolution during standard of care therapies. Clinical progress is restricted due to this limited knowledge of GBM evolution and inability to measure tumor evolution in real-time during therapy. To investigate evolutionary changes during GBM progression, we isolated and profiled GBM sEVs from the plasma of 12 GBM patients with longitudinal blood samples. We subsequently evaluated the relative expression of CNS-related markers ATP1B2, EAAT2, and GSC-related markers CD24, CD44, CD133, and EGFR in sEVs before and during therapy. Furthermore, the expression of ATP1B2, EAAT2, CD44, CD133, and EGFR in primary tumor tissues were confirmed by IHC (n = 11) (Supplementary, Figure S9) to confirm that these markers were expressed in the primary tumor. We then evaluated the results of GEMPAC in relation to clinical interpretation of response to treatments in conjunction with MRI and where available with positron emission tomography (PET) scans.

We explored the capabilities of the GEMPAC score in monitoring disease progression or a positive therapeutic response. Interestingly, when we analyzed the GEMPAC score in patients that had radiological recurrence detected by MRI within 300 days, we identified a significantly increased GEMPAC score at 5-6 weeks, and cycle 5 of TMZ treatment (Figure 5A). Importantly, there was no significant difference in the total abundance of CNS-derived sEVs throughout treatment (Supplementary, Figure S10), highlighting the importance of measuring the total GSC stem cell marker expression to detect therapeutic response. Disease progression in these patients detected by MRI was correlated to a significantly reduced overall survival (Figure 5B), indicating that monitoring patients during therapy with the GEMPAC assay could accurately determine therapeutic response.

**Fig. 5.**
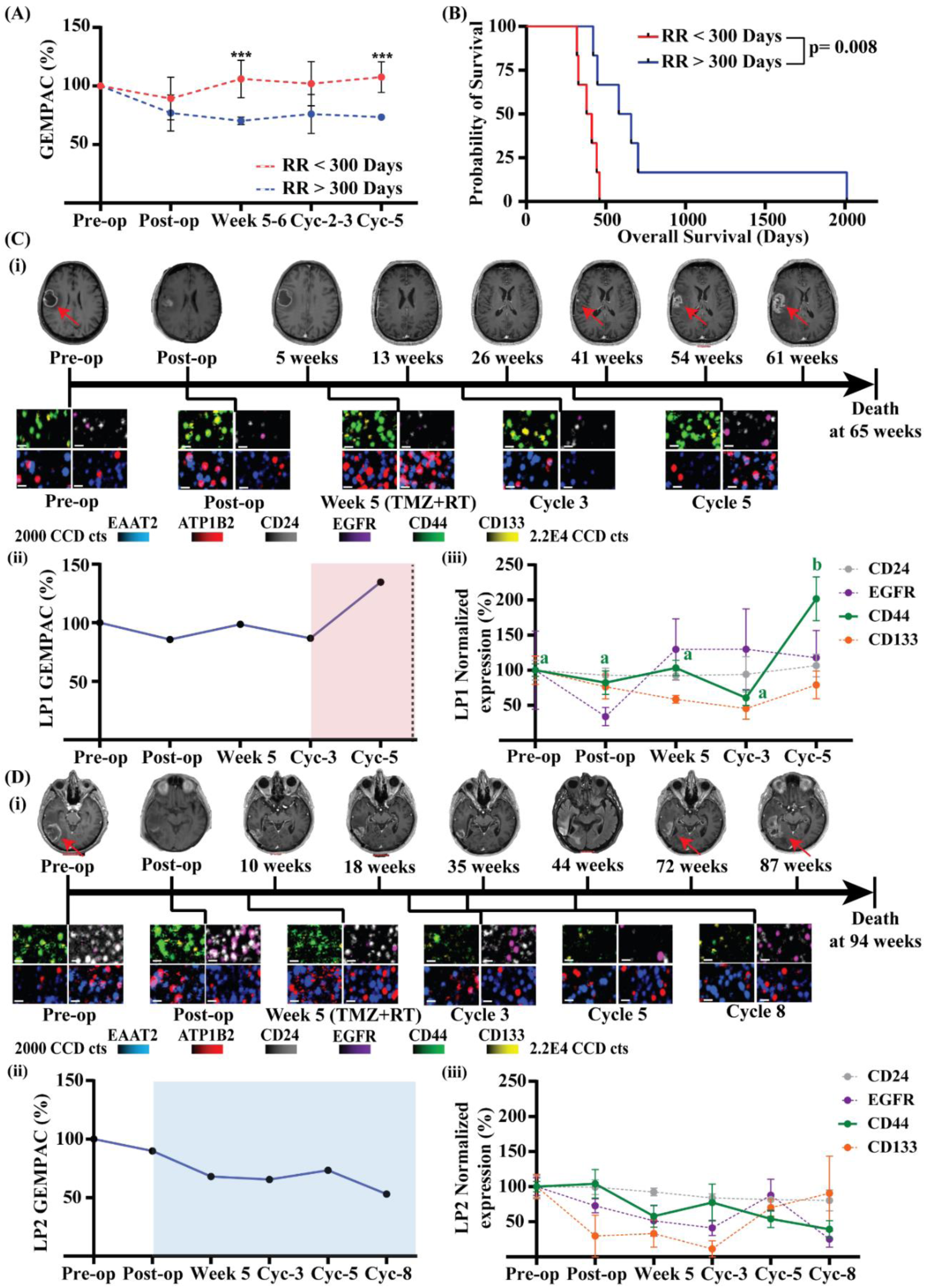
Tracking GSC surface markers on GBM sEVs detects radiological recurrence. (A) The comparison of GEMPAC score between patients that had radiological recurrence (RR) less than 300 days and more than 300 days. Data are represented as mean ± standard deviation. *** p < 0.001. (B) Kaplan-Meier plots of overall survival demonstrate MRI detected radiological recurrence is associated with overall survival. (C) Representative longitudinal case showing detection of progression with the GEMPAC analysis before MRI. (i) MRI images and SERS mapping images of LP1. Red arrows indicated tumor location. Scale bar = 10 µm; (ii) GEMPAC score for LP1 before and during the treatment indicating progression (red area) before MRI detected recurrence at 41 weeks; (iii) Normalized GSC-related protein, CD24 (grey line), CD44 (green line), CD133 (yellow line), and EGFR (purple line) composition changes throughout treatment, indicating a significant increase in the MES-like GSC CD44 markers. (Data are represented as mean ± standard error (n = 4). Significant differences are represented by different letters (p < 0.05). (D) Representative longitudinal case of responding to the TMZ therapy. (i) MRI images and SERS mapping images of LP2. Red arrows indicated tumor location. Scale bar = 10 µm; (ii) GEMPAC score of LP2 before and during the treatment, with the blue area representing a therapeutic response; (iii) Normalized GSC-related protein composition, CD24 (grey line), CD44 (green line), CD133 (yellow line), and EGFR (purple line) changes throughout the treatment, indicating a decrease in the MES-like GSC CD44 marker. Data are represented as mean ± standard error (n = 4).

To further explore and characterize heterogeneity and evolution within malignant cell populations, we assessed the relative GSC protein composition of circulating GBM sEVs at all timepoints (Figure 5C & D; Supplementary, Figure S12). We observed changes in the landscape of GSC-related sEV composition at various stages of disease evolution and during TMZ therapy. Specifically, in representative Longitudinal Patient 1 (LP1), MRI indicated tumor debulking and recurrence, and our GEMPAC assay tracked evolutionary changes of GBM sEVs throughout treatment (Figure 5C (i-iii)). In particular, at cycle 5, an elevated GEMPAC score (Figure 5C(i)) was driven by a subtle increase in the OPC-like GSC CD133 and a significant increase in the MES-like CD44 GSC marker (Figure 5C(iii)) before radiological recurrence was detected at 41 weeks, suggesting disease progression is driven by a clinically relevant glioblastoma stem cell subpopulation consistent with previous studies.[43, 45, 52] Similarly, multiple patients (LP3, 5-7) also exhibited similar evolutionary changes in circulating sEVs with elevated GEMPAC scores being detected in sEVs before MRI indicated tumor progression (Supplementary, Figure S12 & S13).

We next wanted to assess if our GEMPAC assay could monitor positive response to treatment in GBM patients. Our GEMPAC assay detected alterations in the relative expression of CD24, CD44, CD133, and EGFR throughout treatment in representative Longitudinal Patient 2 (LP2) (Figure 5D (i-iii)). LP2 responded positively to TMZ+RT which was recapitulated with our GEMPAC assay showing a dynamic reduction in the GEMPAC score from week 5. This was largely driven by reduced levels of the GSC markers CD44 and EGFR. The response our GEMPAC assay detected was further validated with an overall survival of 94 weeks in this patient. Additionally, our GEMPAC assay consistently measured positive response to treatment with RT and TMZ in additional longitudinal patients LP4, 8 & 9 (Supplementary, Figure S12 & S13).

### Longitudinally monitoring evolution of GSC-related protein composition of GBM sEVs driven by therapy

To directly link the GEMPAC capture and analysis of GBM sEVs to evolutionary changes within the tumor, we compared patients receiving EGFR targeting ABT-414 (depatuxizumab mafodotin) and TMZ+RT (n = 4), versus patients receiving TMZ+RT (n = 5) with matched collection timepoints. Although there was no difference in the GEMPAC score in patients receiving ABT-414 (Supplementary Figure S11), our GEMPAC assay detected evolutionary changes of the tumor through changes in the phenotypic composition of sEVs (Figure 6; Supplementary, Figure S13). In particular, the reduced EGFR expression in a recurrent tumor of a patient treated with ABT-414 compared to TMZ+RT alone (Figure 6A & B), correlated with what we observed in our GEMPAC assay with a significant decrease in EGFR on sEVs (Figure 6C). Due to the molecular and genomic heterogeneity leading to GBM evolution, various subpopulations of cancer cells with stem-like properties following radiotherapy and chemotherapy are capable of driving resistance. In this case, GEMPAC observed a significant increase in the MES-like CD44 GSC marker at cycle 3 and 5 in patients receiving ABT-414 (Figure 6D). Therefore, although ABT-414 suppressed EGFR subpopulations, there was limited therapeutic efficacy against mesenchymal subpopulations.

**Fig. 6.**
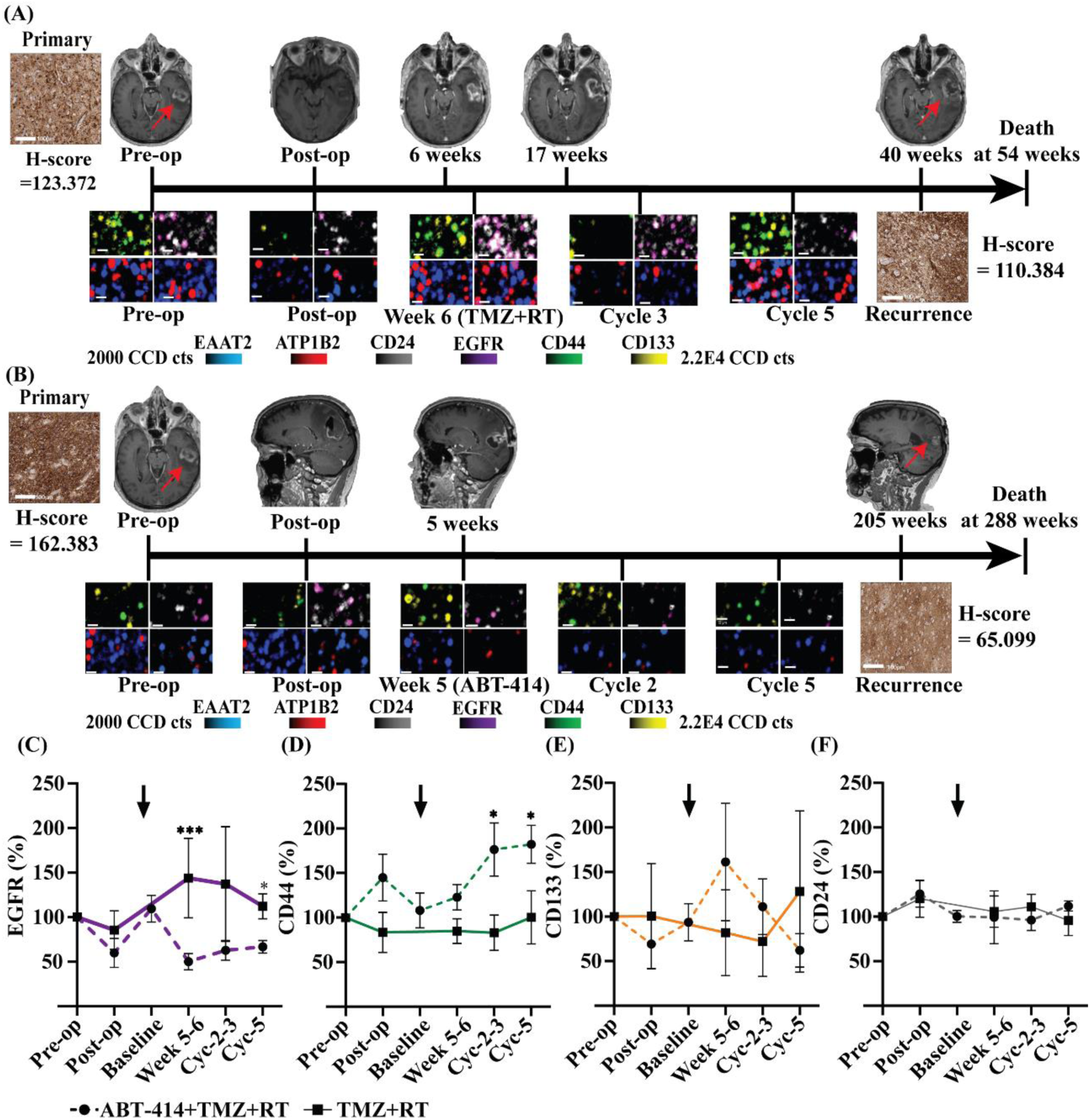
GEMPAC assay tracks tumor evolution by GSC protein composition in circulating GBM sEVs. Representative IHC staining of EGFR in primary and recurrent tumors of patients receiving either TMZ+RT (A, LP3) or ABT-414 with TMZ+RT (B, LP4). Red arrow indicates the tumor location. SERS images scale bar = 10 µm. IHC scale bar = 100 µm. (C-F) Each GSC-related protein expression changes throughout treatment shows significant knockdown of EGFR and increase of CD44 expression in circulating GBM sEVs. * p < 0.05; *** p < 0.001. Solid lines are the patients receiving TMZ+RT (n = 5) and dotted lines are patients receiving ABT-414 with TMZ+RT (n = 4). Data are represented as mean ± standard deviation. Black arrow indicates when the ABT-414 starts.

In addition to our GEMPAC assay measuring treatment efficacy of TMZ+RT, response to the bevacizumab could also be analyzed. Specifically, GEMPAC detected a decrease in GSC markers on circulating sEVs after bevacizumab treatment (Supplementary, Figure S14A). GEMPAC again monitored evolution of the GSC marker composition on GBM sEVs with reduced levels of CD44, CD133 and EGFR at 11 weeks after the start of bevacizumab treatment (Supplementary, Figure S14B & C). Overall, these results demonstrate that GEMPAC has the potential to monitor GBM evolution and therapeutic response in patients being treated with a range of drugs.

## DISCUSSION

In the present study, we developed a nanodiagnostic platform, termed GEMPAC, to provide real-time readout of GBM patients under therapy through analysis of sEVs in blood. A key finding from our work is the identification that sEVs derived from GBM tumor cells can be captured and interrogated in patients. As a result, we were able to monitor the emergence of proliferating GSCs from circulating sEVs to dynamically monitor therapy response and tumor evolution.

The complexity and degree of genomic and cellular heterogeneity of high-grade gliomas such as GBM is only recently being fully comprehended.[53] This is due, in large part, to technological advancements in single-cell profiling leading to numerous discoveries regarding tumor heterogeneity and the revelation of cellular states; specifically MES-like, NPC-like, AC-like, and OPC-like within malignant brain tumors.[31, 54] However, clinical progress in GBM management will be limited without sophisticated approaches to monitor tumor evolution and progression throughout treatment cycle.[55, 56]Advancements in this area have been limited due to difficulties in sampling longitudinal surgical biopsies, and therefore can only be addressed through liquid biopsies in combination with highly sensitive analytical techniques. In addition to the high sensitivity, the GEMPAC has further advantages for liquid biopsy testing in GBM patients as the analysis of sEVs only requires small volumes that can be routinely collected and cryopreserved. This makes sample collection feasible particularly for patients that do not have access to MRI, as they can be monitored regularly regardless of geographical distribution.

At least three of the cellular states within GBM have the capability of propagating tumors in preclinical models, including MES-like, NPC-like and OPC-like.[31–33] Previous studies have proposed quiescent GSCs with MES properties start proliferating in response to therapy and are a driver of disease recurrence.[56, 57] Although the basis for this is unclear, we were able to observe the emergence of a MES-like signature in GBM sEVs by the elevation of CD44 throughout treatment in patients. This elevated level of CD44 on GBM sEVs was associated with disease progression, suggesting that the MES-like GSCs expansion driving recurrent disease[53, 58], can also be monitored dynamically during therapy with the GEMPAC. Another observation in our study was the emergence of elevated CD133 expression on circulating GBM sEVs in a subpopulation of patients, suggesting the expansion of CD133^+^ GSCs. These findings support the premise that CD133^+^ stem cell expansion, which typically have the highest degree of resistance to TMZ [59], persist throughout treatment and could potentially be responsible for the initiation of tumor recurrence.

During our analysis, we did not observe an increase associated with recurrent disease in sEV GSC markers CD24 and EGFR during therapy. Although EGFR is a striking feature of GBM, elevated EGFR expression is associated with tumors with a phenotype dominated by AC-like cells. In terms of EGFR expression in GBM, there is evidence in our data of GBM evolution in response to therapy. In particular, circulating sEVs in patients treated with ABT-414 indicated a shift to a MES-like cellular state with downregulation of EGFR and elevation of CD44, an observation that was seen in the evolutionary changes of the recurrent tumor. There is evidence that a phenotypic shift to a MES phenotype can be driven by multiple factors, including inherent resistance to therapy; a phenotypic shift among individual cells; or changes in the proliferative rates among cell-likes GSC states of GBM.[31, 60] In our study, we were able to observe these changes in real-time as sEVs from MES-like were increasingly abundant following therapy.

Patients often deteriorate rapidly once treatment resistance develops and having an early signal could significantly change treatment by accelerating testing of new drugs, or drug-radiation combinations. Collectively, our GEMPAC platform opens a window for monitoring brain pathologies during therapy and offers a unique view of changes in the GBM sEV landscape at various stages of disease evolution. This assay has potential broad clinical implications and could be utilized to not only predict treatment outcomes, but also determine the phenotypic heterogeneity of GBM. The capability of monitoring the emergence of specific GSCs associated with distinct GBM cellular states might be therapeutically important as targeted elimination of specific GSCs might have clinical benefit. As a result, we envisage with future validation in larger clinical cohorts, the high sensitivity of our GEMPAC platform could be utilized for predicting optimal treatment approaches in new areas of therapeutic development for GBM patients.

## MATERIALS AND METHODS

### Clinical sample acquisition

This study was approved by the Human Research Ethics Committee of the University of Newcastle (H-2020-0231). Informed consent was received from all patients. Blood was obtained from non-fasted patients before surgery, post-surgery (1-7 days) and at the time of treatment in EDTA-coated tubes. Whole blood was centrifuged twice to deplete platelets and separate plasma. Plasma was then aliquoted and stored at −80°C until use. Formalin-fixed paraffin-embedded tumor tissue from surgical resection was sourced from the Mark Hughes Foundation Brain Bank facilitated by the NSW Regional Biospecimen Services for 36 cases of glioblastoma (Supplementary, Table S1). A block containing maximal tumor content was chosen from each patient and diagnosis of glioblastoma confirmed on H&E sections by a neuropathologist (Dr. Ricardo Vilain, Hunter Area Health Service). Clinical information is available in Supplementary, Table S1 including age, gender, treatment information, *MGMT* promoter methylation status, overall survival information, isocitrate dehydrogenase (IDH) status, EGFR status, glial fibrillary acidic protein (GFAP) status, and alpha-thalassemia/mental retardation, X-linked (ATRX) status.

### Bioinformatic analysis to identify brain specific surface proteins

Our objective was to identify robust brain-specific markers for the capture of CNS-specific sEVs. We conducted a bioinformatic analysis to select CNS-specific markers based on data sourced from The Cancer Genome Atlas (TCGA), the Genotype-Tissue Expression (GTEx), and the UniProt database as previously described.[61] This integration enabled a comprehensive investigation of markers specific to the brain. Subsequently, we utilized The Human Protein Atlas, to refine the selection of markers with an extracellular domain and selected candidates with medium to strong immunohistochemistry staining. Following this, the surface markers ATP1B2 and EAAT2 were selected based on the availability of suitable antibodies that target the extracellular domain of each protein.

### Cell culture

Primary patient-derived Q-Cell GBM cell lines were obtained under MTA from QIMR Berghofer Medical Research Institute.[49, 62, 63] BAH1 (QIMR-B001), WK1 (QIMR-B012), and FPW1 (QIMR-B002) were cultured in serum-free KnockOut™ DMEM/F-12 medium supplemented with StemPro® neural supplement (Thermo Fisher Scientific), epidermal growth factor (EGF) (20 ng/mL), and fibroblast growth factor (FGF) (10 ng/mL) (StemPro™ NSC SFM). Cells were maintained as an adherent monolayer on a basement membrane of Matrigel (Corning).[49] For sEV isolation, BAH1, WK1, and FPW1 cells were cultured using Cytodex-1 microcarriers.

### Preparation of microcarriers

Cytodex-1 microcarriers (Cytiva) were prepared according to the manufacturer’s guidelines. Briefly, dry Cytodex-1 microcarriers were hydrated with PBS (50 ml/g Cytodex-1) for at least 4 hours and washed 2 times with the same volume of PBS before being autoclaved (121 °C, 15 psi for 30 mins). Microcarriers were then coated with Matrigel (1:30) before being washed and equilibrated in StemPro™ NSC SFM.

### Cell line sEV medium collection

BAH1, WK1, and FPW1 cells were seeded at 2×10^5^ cells/mL containing 2 g/L of Cytodex-1 microcarriers in 1/3 of the final volume of StemPro™ NSC SFM with periodic stirring at 37 °C for 3 hours. After 3 hours, the volume of the culture was increased to the final volume and continuous shaking commenced at 90 RPM to keep the microcarriers in suspension. Cells were cultured for 72 hours before the culture medium was collected and centrifuged at 500 ×*g* for 10 min and then filtered through a 0.22 µm filter to remove cell debris and large EVs. The filtered culture media was then concentrated using tangential flow filtration as previously described in detail.[64] Briefly, media was prepared for sEV purification by concentrating with a sterile Minimate 300kDa Omega Membrane (Pall Corporation) Tangential Flow Filtration Capsule to approximately 20 mL with continuous diafiltration (5 diafiltration volumes), and further concentrated with a Centricon Plus-70 100 kDa (Merck) to approximately 500 µL before being purified with SEC.

### sEV purification

sEVs were purified from GBM cell culture medium and patient plasma samples using SEC as previously described in detail.[50, 64] Patient plasma samples was thawed from -80°C and kept on ice. Aliquots of 500 µL were centrifuged at 10,000 × *g* for 10 minutes at 4°C to pellet any remaining cellular debris or larger particles. Concentrated culture medium or clarified patient samples were subsequently applied to Izon qEV Original 70 nm columns (Izon Science) and eluted in filtered phosphate-buffered saline (PBS, Sigma-Aldrich). Following the void volume, the following 1.6 mL of sEV-enriched fractions were concentrated to ≤ 50 µL using Amicon Ultra-4 10 kDa MWCO columns (Merck) at 3,500 × *g* for approximately 45 minutes at 4°C. The concentrated sEV isolates were aliquoted and stored at -80°C.

### Nanoparticle tracking analysis (NTA)

NanoSight N300 (NanoSight Nanoparticle Tracking and Analysis Release Version Build 3.4) was used as previously published [64]. Briefly, the samples were diluted with PBS to a particle range of 50 – 100 particles per frame. Samples were analyzed at 25°C. The camera level was set to provide sufficient contrast to identify particles while minimizing background noise and samples were recorded in triplicate.

### Measurement of sEV Size and Concentration using nanoFCM

Purified sEV samples were analyzed by nanoFCM equipped with 488 and 640 nm lasers.Concentration and fluorescence measurements were calibrated with 250 nm silica nanospheres labelled with fluorochromes at 20 mW laser power, 0.2% SS decay and 1 kPa sampling pressure. The particle size distribution of sEVs was calibrated at 10 mW laser power, 10% SS decay and 1 kPa sampling pressure using a four-modal standard silica nanoparticle cocktail (68∼155 nm) which has a similar refractive index to EVs.[65] Positive particles were gated based on a negative control of PBS or the respective fluorescent antibody to adjust for background fluorescence

### sEV immunofluorescent staining for nanoFCM

Approximately 1 × 10^9^ total sEV particles were incubated with either anti-ATP1B2 (PA526279, Thermo Fisher Scientific), anti-EAAT2 (NOVNBP120136, Novus), anti-CD9 APC (17-0098-42, Thermo Fisher Scientific), anti-CD24 (ab134375, Abcam), anti-EGFR (ab231, Abcam), anti-CD44 Alexa Fluor 488 (103016, BioLegend), or anti-CD133 Alexa Fluor 488 (FAB11331G, R&D Systems) in 50 µL of PBS, for 1 h at 37 °C. Labeled sEVs were diluted with 1 mL of PBS and ultracentrifuged at 110,000 × *g* for 30 min using an Optima MAX-XP Benchtop Ultracentrifuge (Beckman-Coulter) TLA-100.3 rotor (k-Factor 14). Samples labeled with anti-ATP1B2 and anti-EAAT2, anti-EGFR, and anti-CD24 were subsequently incubated with goat anti-rabbit IgG H&L APC (ab130805, Abcam), anti-rat IgG2a heavy chain Alexa Fluor 488 (ab172332, Abcam), and anti-mouse IgG Alexa Fluor 488 (ab150113, Abcam) respectively for 1 h at 37 °C, and ultracentrifuged as before. The supernatant was removed and the sEV pellet was resuspended in 50 µL of filtered PBS for analysis with the nanoFCM equipped with 488 and 640 nm lasers. Data was analyzed using FlowJo version 10.8.1.

### Western blot

GBM patient-derived cells (BAH1, WK1, and FPW1) and their sEVs were reduced and denatured by Laemmli buffer and 2-mercaptoethanol (Bio-Rad) for 5 minutes at 95°C. Proteins were resolved by SDS-polyacrylamide gel electrophoresis (SDS-PAGE) and then transferred to polyvinylidene fluoride membranes (Bio-Rad). The membranes were blocked with 5% skim milk in PBS-T (0.1% Tween-20) for 30 minutes at room temperature (RT). Primary antibodies, anti-Calnexin (2679, Cell Signaling) and anti-Hsp70 (610608, BD Biosciences), were applied at the recommended dilutions for 1 hour of incubation at RT. After incubation, the membranes were washed five times with PBS-T and incubated with a HRP-conjugated secondary antibody for 1 hour at RT. The membranes were then washed five times with PBS-T and visualized using enhanced chemiluminescence reagent (Clarity Western ECL Substrate, Bio-Rad) and the Bio-Rad ChemiDoc Imaging System.

### TEM

For TEM analysis, 2.5 µL of isolated sEVs (1 × 10^11^ particles/mL) were fixed with an equal volume of 2% glutaraldehyde for 30 minutes at RT. Subsequently, 5 µL of fixed sample was loaded on Formvar/carbon-coated electron microscopic grids (Electron Microscopy Science) and incubated for 15 minutes and excess liquid was removed by blotting. The grid was washed three times by brief contact with 100 µL of Milli-Q water, followed by blotting to remove excess liquid. To contrast the sample, the grid was negatively stained with 30 µL of 2% uranyl acetate (w/v) for 5 minutes and excess fluid was removed by blotting gently. Grids were left to air dry and observed using transmission electron microscopy (Hitachi HT7700) at 100 kV.

### GEMPAC Chip fabrication

The GEMPAC device with 28 asymmetric electrodes was fabricated with standard photolithography as previously reported.[66, 67] The electrode pattern was designed using Layout Editor L-Edit V15 (Tanner Research) and written on 5-inch soda lime chrome masks (Shenzhen Qingyi Precision Mask Making) using a direct write system μPG 101 (Heidelberg Instruments). Then, a clean 4-inch Boroflat wafer (Bonda Technology Pte Ltd) was spin-coated with a negative photoresist AZnLOF 2020 (Microchemicals GmbH) at 3000 rpm for 30 seconds. After a soft bake for 2 minutes at 110 °C, the wafer was UV-exposed with the above prepared mask at a constant dose of 200 mJ cm^−1^ using a mask aligner (EVG 620, EV Group, St Florian am Inn), following a post-exposure bake for 1 minute at 110 °C and wafer development for 30 s using an AZ726 MIF Developer (Microchemicals GmbH). The gold electrodes were then created by deposition of 10 nm Titanium and 200 nm gold using a Temescal FC-2000 Deposition System (Ferrotec) and overnight lift-off in Remover PG (Microchemicals GmbH). The wafer carrying the gold electrode structures was rinsed with isopropanol and dried under a flow of nitrogen.

To accommodate the liquid sample analysis, a 4 mm-thick polydimethylsiloxane (PDMS) slab with microfluidic well structures was manually aligned to electrodes of the device. The PDMS slab was prepared by curing activated silicon elastomer solution (Sylgard 184, Dow) for 2 h at 65 °C. The PDMS slab was then punched with 5 mm-diameter wells and thermally bonded to the device overnight at 65 °C.

### SERS nanotag synthesis and functionalization

60 nm gold nanoparticles were synthesized through citrate reduction of gold (III) chloride trihydrate (HAuCl_4,_ Sigma-Aldrich).[68] 0.01% (w/v) of HAuCl_4_ was added into 100 mL Milli-Q water and heated till boiling. 1 mL of 1% (w/v) of trisodium citrate dehydrate (Univar Solutions) was quickly added into the boiling solution with constant mixing and kept at a boil for 20 minutes with constant mixing. To synthesize SERS nanotags, 1 mL of gold nanoparticle solution was incubated with 2 µL of 1 mM Dithiobis (succinimidyl pro) (DSP, Thermo Fisher Scientific) and 10 µL of a 1 mM Raman reporter (Sigma-Aldrich) for 5-7 h with gentle shaking (350 rpm) at RT. After that, the solution was centrifuged at 5400 ×*g* for 10 min and resuspend in 200 µL of 0.1 mM PBS buffer. The Raman intensity for each SERS nanotag was measured. After that, the solution was then incubated with 1 µg of paired antibodies, anti-ATP1B2 (PA526279, Thermo Fisher Scientific), anti-EAAT2 (NOVNBP120136, Novus Biologicals), anti-CD24 (ab134375, Abcam), anti-EGFR (ab231, Abcam), anti-CD133 (130108062, Miltenyi Biotec) and anti-CD44 (14044182, Thermo Fisher Scientific) for 30 minutes at RT. The solution was then centrifuged at 600 ×*g* for 6 minutes to remove excess antibodies. After that, SERS nanotags were resuspended in 0.1% (w/v) bovine serum albumin (BSA) to block non-specific binding. The patterns of Raman reporters with antibodies were DTNB-ATP1B2, MBA-EAAT2, TFMBA-CD44, and MPY-CD133.

### GEMPAC functionalization and operation

20 µL of 5 mM DSP were incubated at the center circular electrode for 30 min at RT. Following this, electrodes were washed once with absolute EtOH followed by 3 washes of 1 × PBS. Next, 20 µL of 10 µg/mL of capture antibodies for 2 h at RT were incubated on the electrode. The electrode was then blocked with 5% (w/v) BSA in PBS and incubated overnight at 4 °C. After blocking, electrodes were washed 3 times with 1% (w/v) BSA in PBS before sample addition. A total of 5 × 10^8^ sEVs in 50 µL was added into the circular electrode and an alternative current field of 800 mV and 599 Hz was applied for 45 minutes. Electrodes were then washed 3 times with 1% (w/v) BSA in PBS before the addition of 20 µL of SERS nanotags (1 in 25 dilutions in 1% (w/v) BSA in PBS). The electric field with same conditions as above was carried out for 20 min. Electrodes were then finally washed 3 times with 1% (w/v) BSA in PBS.

### SERS mapping and analysis

Samples were analyzed and recorded by Witec alpha 300 R micro-spectrometer using a 632.8 nm excitation wavelength from a HeNe laser (laser power 4 mW). The integration time for measurement is 0.05 s on each electrode. Each sample has 2-3 separate electrodes as technique replicates. Each electrode was measured with two areas of 60 µm × 60 µm (60 pixels × 60 pixels) squares using a 20 × objective. The signal peak was based on each Raman reporter’s signature peak, MPY – 1000 cm^-1^, MBA – 1078 cm^-1^, DTNB – 1333 cm^-1^, and TFMBA – 1378 cm^-1^. The signal intensity was calculated and representative of each protein marker’s expression level on sEVs.

### Immunohistochemistry

Formalin-fixed paraffin-embedded tumor tissue was sliced into 4 µm full face sections and processed for 3’,3’-diaminobenzidine (DAB) immunohistochemistry using a Ventana Discovery Ultra (Roche) by the HCB. 31 out of 32 patients’ tissues were available for IHC staining. Sections were labeled with rabbit antibodies directed at either EAAT2 ATP1B2, EGFR, CD133 and CD44 (Supplementary, Table S2). All steps, from baking to chromogen addition were performed automatically by the instrument. Tissue sections were baked to slides and deparaffinized, and antigen retrieval then occurred at 95 °C / pH 9 with a total incubation time of 24 minutes before the addition of the primary antibody. The addition of the primary antibody was followed by a 32- minute incubation at 36 °C. Slides were then incubated with a secondary antibody – Anti-Rabbit HQ (Roche), for 20 minutes at 36 °C. Positive control and negative control tissues were included in each batch of slides to confirm the specificity of antibody labeling.

### Quantitative IHC analyses

GBM tissue slides were digitally scanned using the Aperio™ Digital AT2 Pathology System (Leica Biosystems) at 40 × absolute resolution. Quantitative IHC analyses were performed using the HALO™ image analysis platform (version 3.0, Indica Labs, NM). Tissue classification algorithms were used to differentiate tissue regions, such as tumors, necrosis, adjacent normal brain, followed by quantification of pixel intensity values corresponding to DAB staining of protein biomarkers. Area quantification algorithm, which detects biomarker expression across the whole tumor, as well as the cytonuclear algorithm, which quantifies based on cellular compartment, were both used dependent on protein biomarker localization. The labeling intensity of each marker was measured across four representative areas of each tumor and average H-scores for each biomarker for each tumor was calculated as: H-score = (1 x % weak positive tissue) + (2x % moderate positive tissue) + (3 x % strong positive tissue) with a range of 0 - 300. H-scores were then used to create distribution plots to show biomarker intensity across the cohort of glioblastoma cases.

### Normalization of GSC markers and GEMPAC score

CD44, CD133, EGFR and CD24 were normalized by the sum of CNS markers ATP1B2 and EAAT2 at each time point to investigate changes in relative GSC marker expression. To track the abundance of GBM sEVs in circulation, the GEMPAC score is equal to the sum of normalized all GSC markers.

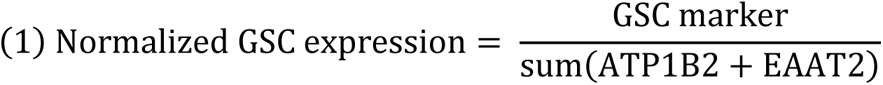

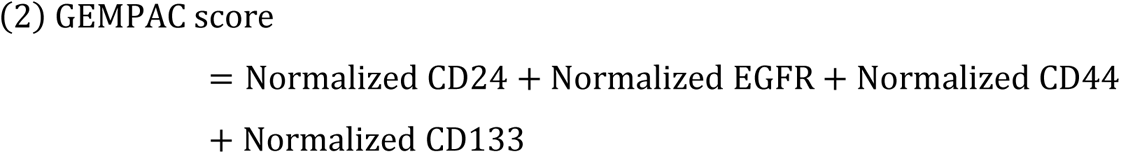

### Statistical analysis

No statistical methods were used to predetermine sample size. All statistical analyses were performed using GraphPad Prism (GraphPad Software v.10.1.2) and values are given as mean ± sem or sd as indicated. When two groups were compared, significance was determined using an unpaired two-tailed t-test. A log-rank test was used to assess significance in Kaplan– Meier survival analysis. One-way ANOVA, or Two-way ANOVA were used for multiple comparisons, and P values adjusted using Tukey, or Šidák for multiple comparisons where it was appropriate. P value threshold of 0.05 was considered statistically significant.

## Data Availability

All data produced in the present study are available upon reasonable request to the authors

## List of Supplementary Materials

Fig. S1 to S14 for multiple supplementary figures

Table S1 to S2 for multiple supplementary tables

## Acknowledgments

The authors acknowledge the staff of the NSW Regional Biospecimen Services, University of Newcastle for sourcing the glioblastoma tumor tissue, bloods and clinical data from the Mark Hughes Foundation Biobank and for completing the immunohistochemistry. We also acknowledge the staff from the Histology Facility at the Hunter Medical Research Institute for cutting the tissue blocks and scanning all the slides. We thank the Mark Hughes Foundation for providing funding for the Biobank and for directly funding this work. X.N. acknowledges the China Scholarship Council (CSC) scholarship.

## Author contribtions

Z.Z., R.J.L., designed and performed experiments, analyzed and interpreted the data, and directed the research. R.L., S.F., P.T., and M.F. were involved in patient sample analysis. Q.Z., was involved in fabrication and chip design. X.N was involved in EV characterization. B.W.D. contributed patient-derived GBM cell lines and supervised cell line study. Z.Z., R.J.L., J.W., S.P., S.R., P.T., and M.T. conceived the study and initiated the research. All authors discussed the results and participated in writing, and revising the manuscript, and approved the submitted version.

## Data and materials availability

The data that support the findings of this study are available from the corresponding author upon reasonable request.

